# Retrospective Cohort Analysis of Nutritional and Respiratory Status in Children with Type III Esophageal Atresia

**DOI:** 10.1101/2025.04.02.25325103

**Authors:** Marie-Alix Chansou, Rony Sfeir, Arnaud Bonnard, Véronique Rousseau, Thomas Gelas, Audrey Guinot, Edouard Habonimana, Pascale Micheau, Aline Ranke, Isabelle Talon, Sabine Irtan, Thierry Lamireau, Pierre-Yves Rabattu, Frédéric Elbaz, Nicolas Kalfa, Nicoleta Panait, Virginie Fouquet, Hubert Lardy, Aurélien Scalabre, Philippe Buisson, Marc Margaryan, Frédéric Auber, Céline Grosos, Corinne Borderon, Cécilia Tölg, Jeanne Goulin, Guillaume Podevin, Frédéric Gottrand, Françoise Schmitt

## Abstract

**Objectives:** To evaluate the impact of undernutrition in school-aged children born with type III esophageal atresia (EA), and to determine its potential risk factors, including their respiratory history and status assessed by pulmonary function tests.

**Methods:** Retrospective multicentre cohort study encompassing patients born between 2008 and 2013 with type III EA included in a national registry. Baseline data, surgical history and outcomes of patients with or without undernutrition (body mass index (BMI) z-score < -2 SD) at the age of 6-9 years were compared.

**Results:** Of the 212 patients included in the study, 20 (9.4%) presented with undernutrition, with a mean BMI z-score of -2.5 +/- 0.4. At birth, 13 (65%) of them where preterm, twice as high as in the control group (34.9%, p = 0.013), but adjusted neonatal weights and associated malformations did not differ between groups. Surgical management of EA and other intestinal malformations, including gastrostomy and fundoplication, were comparable between groups, except for hernia/cryptorchidism occurrence (20% vs 5.2%, p = 0.03). On spirometry, 15 (75%) of these patients demonstrated restriction, as compared to 38% of normal weight patients (p=0.002), and 60% of them required pulmonary treatments (vs 32%, p=0.02). Multivariate analysis identified birth in a level 3 maternity (odds ratio OR=6.0), hernia/cryptorchidism surgery (OR=5.2), a restrictive syndrome (OR=3.3) and pulmonary crisis treatment use (OR=2.7) as risk factors for undernutrition.

**Conclusions:** In contrast to intestinal and esophageal surgeries, the respiratory status appears to be significantly associated with nutritional outcomes in children born with type III EA.

**Clinical trial:** NCT04136795.

**What is known:**

- Undernutrition remains common in children operated on for esophageal atresia.
- There are associations between prematurity and undernutrition in children with esophageal atresia.

**What is new:**

- Undernutrition is associated with a restrictive ventilatory pattern and with the use of pulmonary crisis treatments in school-aged children with type III esophageal atresia;
- On the contrary, in this population, associated malformative conditions including the digestive tract and esophageal surgeries secondary to esophageal repair do not predispose children to undernutrition.

## 1. Introduction

Esophageal atresia (EA) is a rare congenital malformation of the esophagus with an incidence of approximately 1.9 per 10,000 births (1), resulting in two separate segments associated with a caudal tracheoesophageal fistula in its most common form (type III EA (85-90%) of the Ladd classification) (2). Current surgical techniques to restore esophageal continuity and medical advances have resulted in high survival rates, but children can develop significant long-term morbidity, necessitating lifelong multidisciplinary follow-ups (1). The latter should include periodic pulmonary function tests (PFT) and digestive endoscopies during childhood (3,4). Respiratory impairment is thought to occur in approximately 60% of the patients (5) with 30 to 50% obstructive syndrome and 11 to 41% restrictive syndrome (6,7), while digestive complications occur in more than 1/3 of patients with symptomatic gastroesophageal reflux disease (GERD) (4) and 50-85% with dysphagia even into adulthood (8).

Both respiratory and digestive impairments can have a major impact on nutritional status. Several studies have reported rates of undernutrition ranging from 10% to more than 50% during childhood (9,10), and persisting in around 20% of adult patients (8). However, the relationship between nutritional status and respiratory morbidity remains under-explored in the literature. Consequently, the aim of this study was to investigate the potential association between undernutrition and respiratory morbidity in a nationwide cohort of school-age children (6-9 years) operated for type III EA and followed up according to national standardized guidelines, and to identify potential patient- or medical-related factors that might contribute to undernutrition.

## 2. Methods

### 2.1. Patients and setting

A retrospective, national, non-interventional multicenter study was conducted in twenty-eight pediatric surgical centers (ref article 1) in accordance with the principles of the Declaration of Helsinki. The study obtained approval from the Institutional review board (n° 2019/02, 2019/01/16) from the University Hospital Center of Angers. Additionally, it received authorization from the French data protection authority (CNIL ar190017v0) and was declared on clinicaltrials.gov (NCT04136795). Patients’ legal representatives provided informed consent for their inclusion in the study. Eligible patients were selected from the national registry of patients born with EA maintained by CRACMO (Centre de Référence des Affections Congénitales et Malformatives de l’Oesophage, Lille, France)(11). All patients with type III EA born between January 1, 2008 and December 31, 2013 were included in this study. Exclusion criteria were children lost to follow-up or died before the age of four, children with a long gap EA and patients without PFT.

### 2.2. Data recording

Perinatal data collected included antenatal diagnosis of EA, gestational age and birth weight, sex, “inborn” birth in a level 3 neonatal intensive care unit (NICU) hospital, and presence of associated malformations. Low weight for gestational age (LWGA) was defined as a birth weight below the 10th percentile for gestational age (AUDIPOG curves) (12). Surgical data included age at EA surgery, surgical approach and potential concomitant procedures (i.e., fundoplication, anorectal malformation repair, cardiac surgery). Postoperative data included chest tube use, postoperative use of proton pump inhibitors, duration of invasive ventilation, and age at discharge. Any subsequent surgical procedures, whether related to the atresia or not, were also recorded, as were weight, height, and body mass index (BMI) at the 6- and 12-month follow-up visits.

Respiratory, nutritional and digestive monitoring data were collected during the recommended multidisciplinary follow-up consultation between 6 and 9 years (13). The data set included weight, height, BMI and BMI z-score calculation (https://banco.podia.com/calculette-imc-z-score) according to the standard curves of the French Association of Ambulatory Pediatrics (14). Obesity was defined according to the recommendations of the International Obesity Task Force (15). Undernutrition was defined as a BMI z-score below - 2 SD according to the latest French recommendations for the diagnosis of undernutrition in children (16). Digestive disorders including GERD, respiratory history and background treatments, and paraclinical investigations including esophageal dilation, bronchial fibroscopy, chest radiographs, barium contrast study and 24-hour pHmetry were also recorded.

The pulmonary function tests (PFT) data collected were Functional Vital Capacity (FVC), Forced Expiratory Volume in one second (FEV1), the FEV1/FVC ratio, and the calculation of the respective z-scores according to the 2012 Global Lung function Initiative (GLI) international recommendations (17), using software (https://gli-calculator.ersnet.org/index.html). A restrictive syndrome was defined as a FVC z-score < - 1.64, an obstructive syndrome as a FEV1/FVC z-score < - 1.64 and a mixed pattern by both FVC and FEV1/FVC z-scores below -1.64.

### 2.3. Statistical analysis

Statistical analysis was conducted using IBM-SPSS 29.0.0.0 and GraphPad Prism 8.0.2 for Windows software. All tests were two-sided and the statistical level of significance was set at p < 0.05. Patients’ characteristics were described as median with interquartile range or mean +/-standard deviation for continuous variables and as percentages for qualitative variables, based on the available data analysis. When necessary, missing data are indicated in the tables. A comparative analysis between groups of patients with a normal weight or undernourished was performed using the Student t test or a non-parametric Mann-Whitney test for quantitative variables depending on the normality of the distribution (Shapiro-Wilk normality test), and using the Fisher’s exact test for qualitative variables. Factors associated with the occurrence of undernutrition were determined using logistic regression. First, bivariate analysis was employed to select relevant variables, and variables with a p-value below 0.2 entered in the final model. A backward stepwise likelihood-ratio test was used for multivariate analysis, and data were expressed as odds ratios (OR) with 95% confidence intervals.

## 3. Results

### 3.1. Baseline and neonatal care characteristics

Between 2008 and 2013, 590 children underwent surgery for type III EA. Of these, data from 503 patients were analyzable and 256 (43%) patients aged 6 – 9 years had had PFT (Figure 1). After exclusion of patients with missing anthropometric data, 212 patients were finally included in the statistical analysis, of whom twenty (9.5%) were classified as “undernourished status” (US group), while the remaining 192 were of normal nutritional status (NNS group). The neonatal characteristics of the 212 patients are reported in Table 1. Patients of the US group exhibited significantly lower birth weights (2005 [1469 – 2736] g vs 2700 [2135 – 3125] g, p = 0.003) and a higher prevalence of prematurity (65% vs 34.9% p = 0.01). They were also less likely to be born directly in a level 3 NICU hospital (“inborn”) (21.7% vs 10%, p = 0.007). There was no significant difference in the degree of prematurity, prenatal diagnosis or association with VACTERL syndrome, or associated cardiac, renal or digestive malformations.

**Table 1:** Neonatal characteristic of the children and comparison of two groups according to their nutritional status at school age.

|  | Whole cohort<br>(n = 212) | Normal nutritional<br>status<br>(n = 192) | Undernourished<br>status<br>(n = 20) | p-value |
| --- | --- | --- | --- | --- |
| **Baseline characteristics of the patients** |  |  |  |  |
| Birth weight (g) | 2680 [2084 - 3070] | 2700 [2135 - 3125] | 2005 [1469 - 2736] | 0.0025 |
| Low weight for gestational age | 68 (32.1%) | 61 (31.8%) | 7 (35%) | 0.8 |
| Gestational age (WA) | 38.0 [35.0 – 40.0] | 38.0 [35.7 – 40.0] | 35.7 [32.7 – 38.3] | 0.006 |
| Premature birth | 80 (37.7%) | 67 (34.9%) | 13 (65%) | 0.013 |
| Twinning | 18 (8.5%) | 18 (9.4%) | 0 | 0.23 |
| Sex : Boys/Girls | 138 /74 | 123 / 69 | 15 /5 | 0.46 |
| EA antenatal diagnosis | 18 (8.5%) | 16 (8.3%) | 2 (10%) | 0.68 |
| Inborn | 130 (61.3%) | 112 (58.3%) | 18 (90%) | 0.007 |
| VACTERL sequence | 49 (23.1%) | 44 (22.9%) | 5 (25%) | 0.9 |
| Anal malformation | 20 (9.4%) | 19 (9.9%) | 1 (5%) | 0.7 |
| Cardiac malformation | 43 (20.3%) | 36 (18.8%) | 7 (35%) | 0.14 |
| **Esophageal atresia surgical repair and neonatal care** |  |  |  |  |
| Delayed restoration of esophageal continuity | 11 (5.2%) | 8 (4.2%) | 3 (15%) | 0.07 |
| Gastrostomy insertion | 14 (6.6%) | 10 (5.2%) | 4 (20%) | 0.03 |
| Associated surgery | 20 (9.4%) | 19 (10%) | 1 (5%) | 0.7 |
| - Cardiac | 1 (0.5%) | 1 (0.5%) | 0 (0%) | 1 |
| - Anorectal malformation | 21 (9.9%) | 19 (10.0%) | 2 (10%) | 1 |
| - Duodenal atresia | 8 (3.8%) | 8 (4.2%) | 0 (0%) | 1 |
| - Other | 6 (2.8%) | 5 (2.6%) | 1 (5%) | 0.45 |
| Length of invasive ventilation (days) | 3 [1 - 5] | 3 [1 - 5] | 4 [2 - 8] | 0.026 |
| GERD during the neonatal period | 68 (32.1%) | 60 (31.3%) | 8 (40%) | 0.46 |
| Postoperative PPI treatment | 188 (88.7%) | 169 (88.0%) | 19 (95%) | 0.7 |
| Age at discharge from hospital (days) | 21 [15 - 48] | 21 [15 - 45] | 49 [15 - 86] | 0.04 |

Quantitative variables are expressed as median with interquartile range, and qualitative ones
as number (percentages). WA = weeks of amenorrhea; EA: esophageal atresia; Inborn: birth in the level 3 NICU (neonatal intensive care unit) hospital; VACTERL sequence: defined for patient presenting at least two of the following malformations in addition to esophageal atresia: vertebral abnormality, anorectal malformation, cardiac malformation, uropathy, limbs abnormality; PPI: proton pump inhibitors; GERD: gastro-esophageal reflux disease.

**Figure 1:**
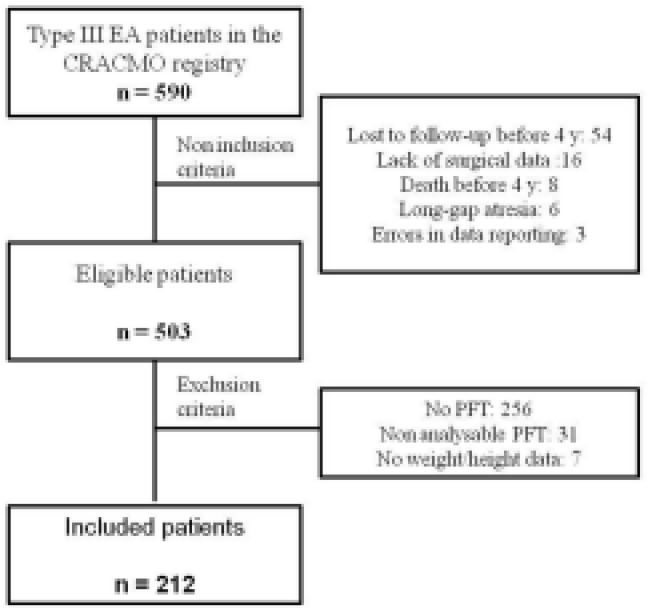
Flow chart of the children with type III esophageal atresia included in the study. BMI: Body Mass Index; PFT: Pulmonary Function Tests; EA: esophageal atresia; CRACMO: Centre de Référence des Affections Congénitales et Malformatives de l’œsophage.

Almost all patients underwent surgery for tracheoesophageal fistula closure on the first day of life, which was associated with restoration of esophageal continuity in 94.8% (n = 201) of the cases. The predominant approach was a right posterolateral thoracotomy in 95.8% (n = 203) of the cases, and the thoracoscopy rate was 3.3% (n = 7). Fourteen children (6.6%) required gastrostomy placement and 20 (9.4%) underwent surgery for associated malformations. There was no difference in EA surgery between the US and NNS groups. Patients in the US group were four times more likely to have a gastrostomy during EA repair (20% vs 5%, p = 0.03), but there was no significant difference between groups in neonatal surgery for associated malformations or the presence and treatment of GERD during NICU stay. However, the duration of invasive ventilation was one day longer and the median hospital stay was twice as long (21[15 – 45] vs 49 [15 – 86] days, p = 0.04) in the US group.

### 3.2. Follow-up after discharge

No statistically significant differences were observed between the US and NNS groups with respect to the incidence of complications related to EA repair, including anastomotic leakage, anastomotic stricture requiring or not esophageal dilation (34% of patients), or tracheoesophageal fistula recurrence. Seventeen children underwent other thoracic surgeries, mainly for cardiac defects, and a total of 81 patients underwent 149 abdominal surgeries, as outlined in Table 2. Except for hernia/cryptorchidism surgery, which was four times more frequent in the US group, there was no difference between the groups.

**Table 2:**
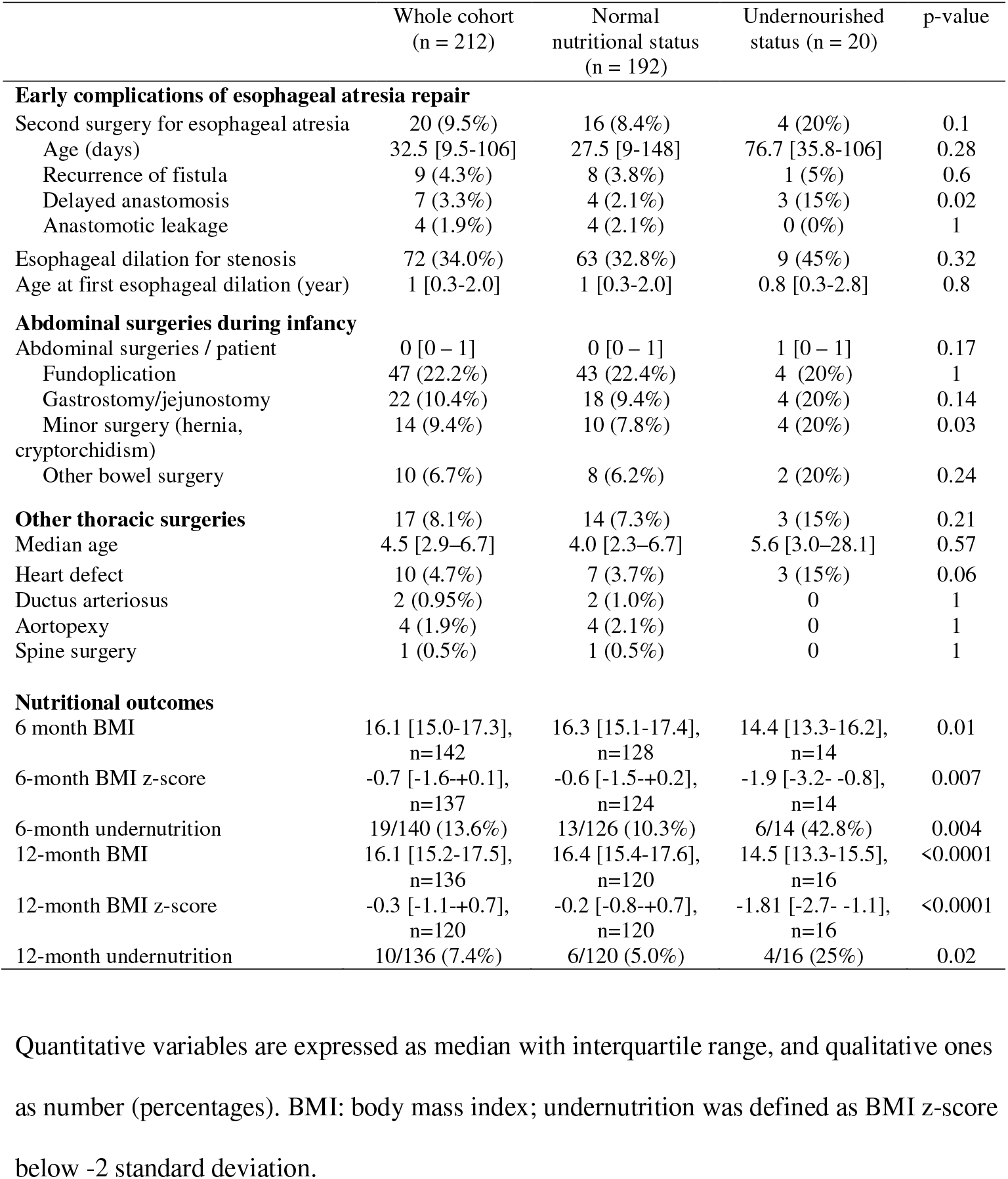
Description of surgical interventions and nutritional outcomes during infancy in the cohort of patients operated on for type III esophageal atresia and comparison of two groups.

There was also no significant difference between the two groups in barium transit and 24-hour pHmetry results at school age (Table 3). Thus, 61 (28.8%) children had undergone at least one barium contrast study at a median age of 6.9 [4.0 – 7.1] years, of which 26 (42.6%) were considered normal and 21.3% and 11.5% of cases respectively exhibiting GERD or stasis. On 24-hour pHmetry, performed at a median of 5.0 [3.0 – 6.6] years, GERD was found in 19 (32.1%) patients. In contrast, pulmonary function tests were abnormal in 90% of the patients in the US group compared to 49.5% in the NNS group (p = 0.0003), with approximately twice as many patients presenting with restrictive syndrome (28.1% versus 55%, p=0.02). Up to 71.7% (152) of the children had a history of respiratory problems and 42.9% (91) of them required chronic pulmonary treatments with no difference between the groups, while the use of crisis treatment in the last six months was 3 times more frequent in the NNS group (32% vs. 10%, p = 0.02).

**Table 3:** Nutritional status, digestive and respiratory history of 6 – 9 years old children with type III esophageal atresia and comparison of two groups according to their nutritional status.

|  | Whole cohort<br>(n = 212) | Normal Nutritional<br>Status<br>(n = 192) | Undernourished<br>status<br>(n = 20) | p-value |
| --- | --- | --- | --- | --- |
| Age (years) | 7.1 [4 - 12] | 7.5 [4 - 12] | 6.5 [6 - 10] | 0.17 |
| **Nutritional and digestive evaluation** |  |  |  |  |
| Weight (kg) | 22.8 [19.0 – 27.0] | 23.3 [20.0 – 27.6] | 17.9 [16.1 – 22.4] | <0.0001 |
| Weight SD | -0.5 [-1.5 - +0.2] | -0.5 [-1.5 - +0.3] | -1.5 [-3.0 - -0.6] | <0.0001 |
| Height (cm) | 122 [115 - 129] | 122 [116 - 129] | 117 [114 - 129] | 0.14 |
| Height SD | -0.5 [-1.0 - +0.2] | -0.5 [-1.0 - +0.3] | -1 [-1.5 - -0.2] | 0.11 |
| BMI (kg/m<sup>2</sup>) | 15.0 [14.0 – 16.6] | 15.2 [14.3 – 16.7] | 12.8 [12.5 – 13.2] | <0.0001 |
| BMI z-score | -0.4 [-1.2 - +0.6] | -0.3 [-1.0 - +0.7] | -2.3 [-2.8 - -2.2] | <0.0001 |
| IOTF classification |  |  |  |  |
| Normal | 161 (75.9%) | 159 (82.8%) | 2 (10%) | <0.0001 |
| Lean | 22 (10.4%) | 4 (2.1%) | 18 (90%) | <0.0001 |
| Overweight/obesity | 21 (9.9%) | 21 (10.9%) | 0 (0%) | 0.23 |
| Clinical GERD | 38 (17.9%) | 36 (18.8%) | 2 (10%) | 0.54 |
| Treatment of GERD | 47 (22.2%) | 44 (22.9%) | 3 (15%) | 0.58 |
| Barium contrast study | 61 (28.8%) | 56 (29.2%) | 5 (25%) | 0.8 |
| Age (year) | 6.9 [4.0 – 7.1] | 7.0 [4.0 – 7.5] | 6.3 [6.0 - 6.8] | 0.64 |
| Normal | 26 (42.6%) | 23 (41.1%) | 3 (60%) | 0.39 |
| GERD | 13 (21.3%) | 13 (23.2%) | 0 (0%) | 0.57 |
| Stasis | 7 (11.5%) | 6 (10.7%) | 1 (20%) | 0.47 |
| 24-hour pHmetry | 56 (26.4%) | 53 (27.6%) | 3 (15%) | 0.29 |
| Age (year) | 5.0 [3.0 – 6.6] | 5.0 [2.9 – 6.5] | 4.0 [2.0 - 7.6] | 0.89 |
| GERD | 19 (32.1%) | 17 (32.1%) | 2 (66.7%) | 0.4 |
| **Pulmonary evaluation** |  |  |  |  |
| Respiratory history | 152 (71.7%) | 136 (70.8%) | 16 (80%) | 0.6 |
| Asthma | 58 (27.4%) | 54 (28.1%) | 4 (20%) | 0.6 |
| Current respiratory treatments |  |  |  |  |
| None | 118 (55.7%) | 108 (56.3%) | 10 (50%) | 0.64 |
| Background treatment | 91 (42.9%) | 81 (42.2%) | 10 (50%) | 0.64 |
| Acute crisis treatment | 75 (35.4%) | 63 (32.0%) | 12 (10%) | 0.02 |
| Pulmonary function tests |  |  |  |  |
| FVC | 1.35 [1.10 – 1.63] | 1.37 [1.13 – 1.66] | 1.13 [0.85 - 1.41] | 0.087 |
| FVC z-score | -0.95 [-1.81 - -0.25] | -0.81 [-1.67 - -0.07] | -1.90 [-2.81 - -0.97] | 0.002 |
| FEV1/FVC | 90.2 [83.6–96.0] | 90.3 [83.7-95.9] | 88.8 [82.1-96.3] | 0.76 |
| FEV1/FVC z-score | 0.07 [-1.02 - +0.99] | 0.08 [-1.02-+0.97] | -0.40 [-1.08-+1.24] | 0.76 |
| Normal | 99 (46.7%) | 97 (50.5%) | 2 (10%) | 0.0003 |
| Pure restrictive pattern | 65 (30.7%) | 54 (28.1%) | 11 (55%) | 0.02 |
| Pure obstructive pattern | 25 (11.8%) | 22 (11.5%) | 3 (15%) | 0.71 |
| Mixed pattern | 23 (10.8%) | 19 (9.9%) | 4 (20%) | 0.24 |

BMI: body mass index; IOTF: International Obesity Task Force; GERD: gastro-esophageal
reflux disease; FVC: Forced Vital Capacity; FEV1: Forced Expiratory Volume in one second.
Quantitative values are expressed as median with interquartile range, and qualitative variables as number and percentages.

### 3.3 Nutritional status and undernutrition-associated factors

Available nutritional status during the first year showed lower median BMI z-scores in the US group at 6 and 12 months (Table 2), with, respectively, 4 times (42% vs 10%, p = 0.0004) and 5 times (25% vs 5%, p = 0.02) more undernourished children. At a median age of 7.1 [4 - 12] years, median weight was 22.8 [19.0 – 27.0] Kg and median BMI was 15.0 [14.0 – 16.6] kg.m^-2^, with a difference of 2.4 points between the US and NNS groups (12.8 vs 15.2 kg.m^-2^, p < 0.0001), leading to significant differences in IOTF classification (Table 3).

Factors associated with undernutrition at school age were identified using the differences between the US and NNS groups identified among qualitative variables in the bivariate analysis. These included patient-related factors (prematurity, heart disease, gastroesophageal reflux) and care-related factors (gastrostomy, thoracic surgery, minor abdominal surgery, restrictive syndrome and use of crisis respiratory treatment). Too many missing data prevented the inclusion of growth parameters at 6 and 12 months in the model. Factors statistically associated with undernutrition were being “inborn” (Odds ratio (OR): 6.033 [1.287 - 28.272], p = 0.023), restrictive syndrome on PFT (OR = 3.344 [1. 225 - 9,174], p = 0.018), minor abdominal surgery (OR = 5,208 [1,276 - 21,277], p = 0.22) and use of respiratory crisis treatment in the past 6 months (OR = 2,740 [0.985 - 7,634], p = 0.053).

## 4. Discussion

In the present study, the prevalence of undernutrition in children, as determined by a BMI z-score below -2 SD, was 9.4%. This result is consistent with data from the literature, despite large variations in the definition of “undernutrition” and the selected undernutrition index, as well as in the age at assessment. For example, previous studies found 41% of 10-year-old children below the 25th weight percentile (18), 4.8% of undernourished children assessed by standard deviations of weight and height (19), or 9% of 1-year-old children with low weight-for-height z-scores (10). Historically, the Waterlow index (weight/expected weight for height) has been the most widely used method to define and grade undernutrition (20), but the superiority of the BMI z-score over other indicators has been validated (21), and its use is currently recommended in children and adults (16). Nevertheless, these anthropometric scores remain an approximation of the patient’s nutritional status, as undernutrition reflects a mismatch between needs and intake (22). Its diagnosis is based on phenotypic criteria (e.g., weight loss, changes in growth curves) and etiological criteria (e.g., reduced intake, malabsorption, acute and chronic pathologies). However, these criteria cannot be taken into account in a retrospective study that lacks data on food intake and changes in weight and height.

In this series of patients operated for type III EA with both anthropometric and respiratory evaluation at school age, four factors appeared to be significantly associated with undernutrition, including birth in a level 3 NICU, history of minor abdominal surgery, presence of a restrictive pattern on PFT and use of respiratory crisis treatment in recent months.

With an odds ratio of 3.3, to our knowledge, this is the first study to demonstrate an association between undernutrition and restrictive syndrome in children with type III EA. In adults with restrictive syndromes, the rate of undernutrition varies between 9% and 55% in different pathologies such as diffuse interstitial lung disease, or restrictive syndromes secondary to morbid obesity (23,24). In children, the relationship between respiratory impairment and nutritional status has been studied in cystic fibrosis (25) or in premature infants (26), particularly those with pulmonary bronchodysplasia (27,28). In premature infants, Turner *et al*. report a reduction in FVC at 10 years of age with no clinical impact on respiratory function (29). The mechanisms of undernutrition in children with chronic respiratory disease are numerous, including anorexia, increased energy expenditure through elevated respiratory workload, superinfections, inflammatory syndrome, and gastric emptying problems (30).

On the other hand, malnutrition may adversely affect lung maturation from fetal life onward (31) and/or induce adaptive mechanisms such as “lung-sparing growth,” which preserves lung growth by maintaining an adequate thoracic size at the expense of a reduction in leg size (32). Other studies have reported a decrease in FVC and FEV1 without change in FEV1/FVC ratio and a decrease in thoracic diameter without functional consequences in malnourished children (33,34).

In multivariate analysis, we also found that patients in the undernourished group were twice as likely to have had treated asthma attacks in the last few months. The association between acute respiratory episodes and nutritional status, as previously reported by Lejeune *et al*. (35), has also been documented in younger infants. Their findings indicated that children with z-score height for age < -2 SD were more prone to hospitalization for respiratory events during the initial year of life.

Another factor strongly associated with undernutrition was being born in a university hospital center. This may be due to the preferential referral of pregnant women with fetuses exhibiting abnormal ultrasounds, intrauterine growth retardation, or maternal pathologies to level 3 maternity units. Children in the undernourished group were about twice as likely to be born prematurely and had 25% lower birth weights, although there was no significant difference in low weight for gestational age. A recent study by Depoortere et al. (36) identified prematurity, low weight for gestational age and VACTERL syndrome as risk factors for undernutrition at one year of age. Similarly, Birketvedt *et al*. (37) found that low birth weight was associated with adolescent malnutrition. In the present series, no associated malformative condition was identified as a risk factor for undernutrition. With regard to surgical data, only minor surgery was significantly associated with undernutrition in this study. This finding has not been previously described and may again be related to prematurity, as inguinal hernia and cryptorchidism are more prevalent in this population (38,39). Interestingly, no other type of surgery, whether derived from EA repair or not, including esophageal dilation, anti-reflux surgery, gastrostomy, anorectal malformation surgery, other bowel surgery, or cardiac surgery, was associated with undernutrition. This is in contrast to some studies describing GERD and fundoplication as predisposing factors for childhood malnutrition (10,19,40).

Limitations of our study include its retrospective design, which introduces information and measurement biases and deficiencies, particularly regarding anthropometric data. Additionally, the multicenter design, while necessary to achieve a sufficiently large cohort, is subject to variability in follow-up between centers. Another weakness is the lack of comprehensive data on patient diet, potential eating disorders, and nutritional support, which are known issues in this population. Finally, the relatively small number of undernourished patients in relation to the entire cohort complicates statistical analysis and precludes the possibility of conducting stratification or multiplicity adjustments.

The strengths of our study lie in the large number of patients, comparable to other published research, and the standardization of esophageal atresia (EA) care and surveys at the national level, ensuring cohort homogeneity. The study employed the most recent validated methods for PFT analysis and undernutrition characterization. However, prospective randomized studies that control for potential confounders such as prematurity are essential to confirm these initial findings.

## 5. Conclusion

The prevalence of undernutrition remains high in children who have undergone surgery for esophageal atresia. Children born prematurely or with low birth weight demonstrate a heightened risk of inadequate nutritional status at school age, on the contrary to those who have had surgery for intestinal or cardiac malformations, fundoplication, or complications from EA repair. Respiratory complications later in life may further increase the risk of malnutrition, particularly in children with restrictive breathing patterns. These findings underscore the need for close monitoring of these children, including regular multidisciplinary consultations that systematically incorporate pulmonary assessments and nutritional interventions to identify and address undernutrition.

## Data Availability

All data produced in the present study are available upon reasonable request to the authors

